# Genomic reconstruction of an azole-resistant *Candida parapsilosis* outbreak and the creation of a multilocus sequence typing scheme: a retrospective observational and genomic epidemiology study

**DOI:** 10.1101/2024.02.22.24302918

**Authors:** Phillip Brassington, Frank-Rainer Klefisch, Barbara Graf, Roland Pfüller, Oliver Kurzai, Grit Walther, Amelia E. Barber

## Abstract

**Background:** Fluconazole-resistant *Candida parapsilosis* has emerged as a significant healthcare-associated pathogen with a propensity to spread patient-to-patient and cause nosocomial outbreaks, similar to *Candida auris*. This study investigates a prolonged outbreak of fluconazole-resistant *C. parapsilosis* across multiple years and healthcare centers in Berlin, Germany.

**Methods:** In this retrospective observational study, we used whole-genome sequencing of isolates from the outbreak in Berlin and other regions within Germany and compared them with isolates from a global distribution to understand the molecular epidemiology of this outbreak. Additionally, we used the genomic dataset of global samples to identify loci with high discriminatory power to establish a multi-locus sequence typing (MLST) strategy for *C. parapsilosis*.

**Findings:** A clonal, azole-resistant strain of *C. parapsilosis* was observed causing 33 cases of invasive infection from 2018-2022 in multiple hospitals within the outbreak city. Whole genome sequencing revealed that outbreak strains were separated by an average of 36 single nucleotide variants, while outbreak strains differed from outgroup samples from Berlin and other regions of Germany by an average of 2,112 variants. Temporal and genomic reconstruction of the outbreak cases indicated that transfer of patients between healthcare facilities was likely responsible for the persistent reimportation of the drug-resistant clone and subsequent person-to-person transmission. German outbreak strains were closely related to strains responsible for an outbreak in Canada and to others isolated in the Middle East and East Asia. Including the outbreak clone, we identified three distinct *ERG11* Y132F azole-resistant lineages in Germany, marking the first description of this azole-resistance in the country and its endemic status. Using the novel MLST strategy, a global collection of 386 isolates was categorized into 62 sequence types, with the outbreak strains all belonging to the same sequence type.

**Interpretation:** This study underscores the emergence of drug resistant fungal pathogens that can spread patient-to-patient within a healthcare system, but also around the globe. This highlights the importance of monitoring *C. parapsilosis* epidemiology globally and of continuous surveillance and rigorous infection control measures at the local scale. Through large-scale genomic epidemiology, our study offers a high-resolution view of how a drug-resistant clone behaved in a local healthcare system and how this clone fits into the global epidemiology of this pathogen. We also demonstrate the utility of the novel typing scheme for genetic epidemiology and outbreak investigations as a faster and less expensive alternative to whole genome sequencing.

**Funding:** German Federal Ministry for Education and Research, German Research Foundation, German Ministry of Health

**Research in context:** *Evidence before this study:* We searched PubMed and Google Scholar from database inception to Apr 25, 2024, using the search terms “Candida parapsilosis”, “outbreak”, “azole resistance”, and/or “fluconazole” in PubMed and Google Scholar. We applied no language or study type restrictions. The epidemiology of candidemia has undergone dramatic changes in recent years. New pathogenic species, such as *Candida auris*, have emerged, and existing species like *Candida parapsilosis* have increased in prominence. There has also been a worrying increase in drug resistance among *Candida* species. Moreover, numerous drug-resistant outbreaks of *C. parapsilosis* have been reported worldwide and are challenging to control due to their prolonged and intermittent nature. The overwhelming majority of previous work has used microsatellite markers to infer genetic relationships among outbreaks strains, obscuring whether they are really clonal in nature, our understanding of the temporal and transmission dynamics of these outbreaks, and the genetic relationship between outbreak clones.

*Added value of this study:* This study adds to the existing evidence by utilizing whole genome sequencing in conjunction with hospital records to analyze a prolonged outbreak of clonal, azole-resistant *C. parapsilosis* that occurred across multiple years and medical centers. This study demonstrates that patient transfers can result in the reimportation of outbreak clones, posting a significant challenge for infection control. We also reveal that the outbreak clone is closely related to drug-resistant isolates from other continents, highlighting the global spread of drug-resistant *C. parapsilosis*. Furthermore, the study addresses the need for rapid strain differentiation in outbreak settings by establishing and validating a set of four loci for Sanger sequence-based typing, which provide a highly discriminatory tool for epidemiologic investigations.

*Implications of all the available evidence:* This study underscores the global challenge of azole-resistant *C. parapsilosis* and its importance as the causative agent of nosocomial outbreaks. Clinicians should be aware of the evolving epidemiology of *C. parapsilosis* and the prevalence of drug-resistant strains, emphasizing the importance of appropriate antifungal stewardship and infection control measures. The study emphasizes the challenges caused by inter-hospital transmission and their role in persistent outbreaks, highlighting the need for robust surveillance and coordination among healthcare facilities. While whole genome sequencing (WGS) is becoming more widely available, it is still not available in many settings due to cost, limitations in bioinformatic expertise, and the absence of standardized methodology and data interpretation. The establishment of a sequence-based typing scheme is a valuable tool for rapid assessment of samples, which can aid in outbreak tracking and containment efforts, and provide results more rapidly even in settings where WGS is available.

## Introduction

During the last decade, the epidemiology of candidemia has seen dramatic changes due to the emergence of new pathogenic species like *Candida auris,* and dramatic increases in the prominence of existing species such as *Candida parapsilosis* ^1–3^. *C. parapsilosis* is now the second to fourth leading cause of systemic *Candida* infections and has gone from being historically associated with pediatric infection to a frequent cause of catheter-associated bloodstream infections in adults ^1^. A worrying accompaniment to the changing species epidemiology is a corresponding increase in drug resistance in *Candida* spp, including *C. parapsilosis*. Previously, isolates of *C. parapsilosis* were overwhelmingly azole susceptible. Now, there are more than 36 studies from 15 countries documenting local fluconazole resistance rates of >10% in *C. parapsilosis* and an exponential increase in reports of fluconazole-resistant *C. parapsilosis* since 2019 ^3^. Fluconazole-resistant invasive disease results in a three-fold increase in mortality compared to infection with a susceptible strain (16% for susceptible to 50% resistant infections) ^4^. The most dominant fluconazole resistance mechanism seen clinically is the Y132F amino acid substitution in the azole target gene, lanosterol 14-alpha-demethylase (*ERG11*).

Alongside the rapid increase in fluconazole resistance, there has been an alarming number of drug-resistant outbreaks of *C. parapsilosis* reported since 2018 ^5–13^. This organism is commonly found on the skin, and, as a result, it can spread within healthcare facilities in a patient-to-patient manner through contaminated medical devices, environmental surfaces, or the hands of healthcare workers ^5,9^. Outbreaks of *C. parapsilosis* can also persist despite strict infection control strategies and may show gaps in the occurrence of outbreak strains ^3,14^. Recognition of nosocomial outbreaks and implementation of successful infection control measures requires a rapid and reproducible method for discriminating closely related isolates. However, there is currently no gold standard for strain typing in *C. parapsilosis*. Currently, the most widely used method is a species-specific set of microsatellite markers ^15^. However, microsatellite typing requires specialized equipment, is prone to PCR artifacts, and requires specialized knowledge for interpretation ^16^. Moreover, microsatellite-based typing results are not as easily shared and compared across centers. Whole genome sequencing (WGS) has exceptional resolution for discriminating between closely related isolates and is incredibly valuable for outbreak investigation, as has been demonstrated for *C. auris* ^17,18^. However, WGS is not feasible for many settings due to its cost, time to result, and limitations in trained personnel to analyze the data. The aim of this study was to investigate a prolonged outbreak of azole-resistant *C. parapsilosis* using WGS to understand its epidemiology and to introduce a sequence typing strategy for rapid and inexpensive outbreak investigations.

## Methods

### Study design, outbreak description, and isolate collection

In this retrospective, national study, the genomes of *C. parapsilosis* isolates sent to the NRZMyk were sequenced. Samples were submitted by German healthcare facilities and laboratories for susceptibility testing between 2016 and 2022. All potential outbreak samples were included in the study based on the following criteria: samples originated from Berlin, samples were resistant to fluconazole (FLC^R^) and voriconazole (VOR^R^), and were susceptible to posaconazole. Non-outbreak isolates were selected for inclusion in the study from the NRZMyk strain collection as follows: all strains that originated from outside Berlin and displayed resistance to one or more azole were included. For the azole resistant samples from outside Berlin, a matched number of azole susceptible strains from the city were randomly selected for inclusion from the NRZMyk collection.

Outbreak cases were initially detected in December 2018 and January 2019 by physicians at Hospital #1 (name anonymized for privacy) (Figure 1a), who noticed an increase in isolates with a distinct antifungal susceptibility profile. The isolates were resistant to fluconazole (FLC^R^) and voriconazole (VOR^R^), but susceptible to other triazoles. After a period of approximately five months, during which no FLC^R^ and VOR^R^ infections were reported at Hospital #1, infection isolates with this susceptibility profile were once again observed (Figure 1a). The outbreak was officially declared at that time (October 2019) and was formally investigated including environmental surveillance. As *C. parapsilosis* is known to colonize the ear, 21 ICU patients at Hospital #1 were screened with swabs of both ears, but none were positive for *C. parapsilosis*. To investigate possible transfer by staff, sixteen bedside stethoscopes were screened. Only one sample was positive, but the resulting *C. parapsilosis* isolate was azole susceptible. To prevent further cases, emphasis was placed on hygiene measures and hand disinfection. Following implementation of the infection control measures there was a nine-month period without FLC^R^ and VOR^R^ isolates reported. However, additional cases were detected in September 2021 and the start-stop pattern continued through 2022 (Figure 1a). Consultations with other hospitals and clinical laboratories in Berlin identified *C. parapsilosis* infections with the same antifungal susceptibility profile at five other institutions (H0, H8, H9, H10, H14). As outbreaks of *C. parapsilosis* are being increasingly reported, there is an urgent need for a rapid and reproducible tool capable of differentiating isolates. Motivated by this, we also set out to identify a minimal set of loci that could be used for Sanger sequence-based typing for both outbreak detection and epidemiology in our study. The Ethics Committee of the University Hospital Jena approved this study under registration number 2024-3255-Daten.

**Figure 1.**
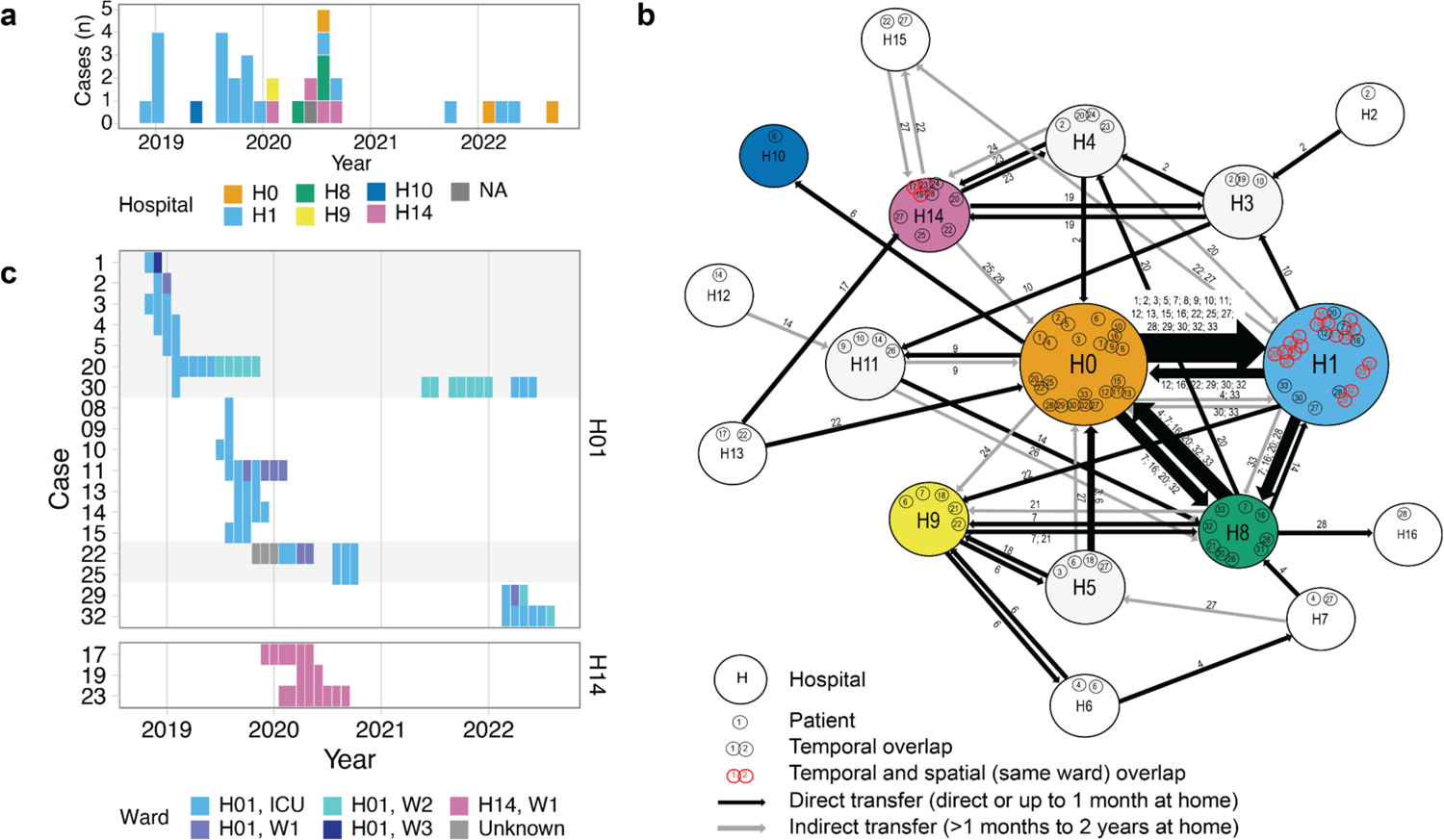
Timeline and hospital network dynamics of a *Candida parapsilosis* outbreak in Berlin, Germany. (a) Timeline histogram of the 33 outbreak cases. Dates are taken from the date of first positive culture. Color indicates the inpatient hospital at the time of isolation. A single case was identified in an ambulatory patient, which is indicated with “NA” for the inpatient hospital. (b) Inpatient stays of the outbreak cases represented as a network, where nodes (large circles) represent hospitals in the Berlin area and edges (lines) represent direct or indirect patient transfers between two institutions. Edge thickness depicts the relative number of transfers between two nodes. Individual patient case numbers are indicated within the node and along the edge. For the network, admission and discharge data beginning 12 months prior to the first detected outbreak and extending until the end of 2022 was utilized. (c) Timeline of 21 cases with temporal and spatial overlap. Alternating gray and white fills indicate five patient clusters where temporal and spatial (same ward) overlaps with other outbreak cases could be verified from inpatient records.

### Procedures

Antifungal susceptibility testing was performed by the NRZMyk using the methodology described in appendix 1 p 2. For tracking outbreak dynamics, admission and discharge records were obtained for all outbreak patients beginning January 2018, corresponding to one year before the first detected outbreak case, and extending to December 2022 and the network of patient transfer manually assembled. These dates correspond to roughly 12 months prior to the first case and extending until there had been several months without new cases in 2022. Patient information, including date of birth, sex, underlying disease, and antifungal treatment history, was obtained from the intake form of the NRZMyk.The NRZMyk does not collect information on patient ethnicity or race.

### Whole genome sequencing and computational analysis

DNA extraction was performed as described in the appendix 1 p 2. Library preparation and paired end sequencing was performed by GeneWiz (Leipzig, Germany) or Novogene (Cambridge, UK) using the Illumina MiSeq platform and NovaSeq platforms, respectively (Illumina, San Diego, CA, USA). One outbreak sample was sequenced a second time using new outgrowth and DNA extraction from the freezer stock to assess the degree of technical variation between sequencing runs. Illumina data of global samples was obtained from the NCBI Sequence Read Archive and was included in all analyses except for those exclusively examining outbreak samples or comparisons between outbreak and other German samples. Sequence data quality control, processing, and identification of variants are described in appendix 1 p 2. The SNV-based phylogeny and all SNV distance-based analyses only considered variant positions with non-zero coverage in all 386 samples. Additional information about phylogeny construction, how genetic distances were calculated, and root-to-tip regression for assessing clock-like evolution are detailed in appendix 1 pp 2-3.

### Sequence typing

The process by which loci were screened and identified for sequence typing and how the number of loci was selected is described in appendix 1 p 3. PCR amplification of typing loci was performed using the primers and cycling parameters described in Table 2. Our Sanger sequence provider did not report heterozygous sites with IUPAC codes and performed background subtraction, which in rare cases erroneously removed a heterozygous peak. These were resolved by requesting the unprocessed chromatograms and manually reviewing them to ensure that IUPAC bases were reported correctly. The samples used for validation were collected from three hospitals in Berlin and were suspected to be additional outbreak cases based on their antifungal susceptibility profile (FLC^R^, VOR^R^, POS^S^) and their isolation from hospitals with previous outbreak cases. Hunter’s discriminatory power was calculated as follows, where N represents the number of strains tested and x_j_ the number of strains belonging to the jth type:

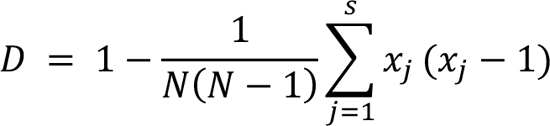

### Statistical analysis

Statistical analyses were performed using R version 4.3.1 and significance was defined as p < 0.05. Mann-Whitney U test was used to determine the pairwise genetic difference between outbreak and outgroup strains. Spearman correlation was used to assess the relationship between isolate date and root-to-tip distance. ANOVA followed by Tukey’s HDS test was used to test for pairwise SNV differences between the outbreak strains and the closely related strains from other countries.

## Results

In this study, we included 38 outbreak isolates from 33 cases identified through epidemiological and genomic investigations. All isolates were fluconazole resistant with a median MIC of 64 mg/L (IQR 16 to >64) and voriconazole (median MIC 1 mg/l; IQR 1.25), but susceptible to posaconazole (Table 1). We also included 38 non-outbreak isolates from the NRZMyk, originating from unlinked hospitals in Berlin and outside Berlin displaying resistance to one or more azole (Table 1). Most outbreak cases were first reported as wound infections (39%; 12/33), followed by infections associated with central venous catheters (29%; 9/33) (Table 1). Antifungal treatment histories were available for 30 cases (appendix 1 p 5). 33% (10/30) of patients were treated with an azole within 60 days of *C. parapsilosis* isolation, while a larger proportion (53%; 16/30) were treated with an echinocandin within the previous 60 days (Table 1). The defined daily dose per 100 bed days of azole antifungals at Hospital #1 was similar to that of other care level three hospitals in the region and remained largely stable over the five-year study period (appendix 1 p 5).

Reconstructing the outbreak transmission dynamics revealed that even though the largest fraction of cases (n = 20) were diagnosed at Hospital #1, most of these patients had been directly transferred to this facility from a major surgical facility (Hospital #0 in Figure 1b). Supporting nosocomial transmission, 94% of outbreak cases (31/33) had verified temporal overlap with other cases. Ward information was available for 28 cases at hospitals H1 and H14, revealing five patient clusters of 21 patients with spatial and temporal overlap on the same ward at the same time as other outbreak cases, strongly supporting patient-to-patient spread (Figure 1c). Altogether, we report a multiyear, sporadic outbreak of azole-resistant *C. parapsilosis* in Germany that was not limited to any single hospital but involved a network of hospitals in Berlin.

To examine the genomic relationship between the included isolates, we performed whole genome sequencing (WGS). 68% (51/75) of the sequenced samples originated from Berlin, including 38 isolates from the 33 cases identified to be part of the outbreak and 13 non-linked isolates from different medical centers in Berlin. Publicly available data for an additional 311 samples was included to facilitate comparison with the global *C. parapsilosis* population, bringing the total number to 386 genomes. WGS samples analyzed had a mean depth of coverage of 123x±80 (SD) (appendix 2).

When built into a phylogeny, we did not observe a strong population structure among the 386 samples and no lineage was unique to a particular country or continent (Figure 2). The close genetic relationship between the outbreak isolates was evident by their tight grouping and short branch lengths, supporting the nosocomial transmission that was inferred from the outbreak reconstruction.

**Figure 2.**
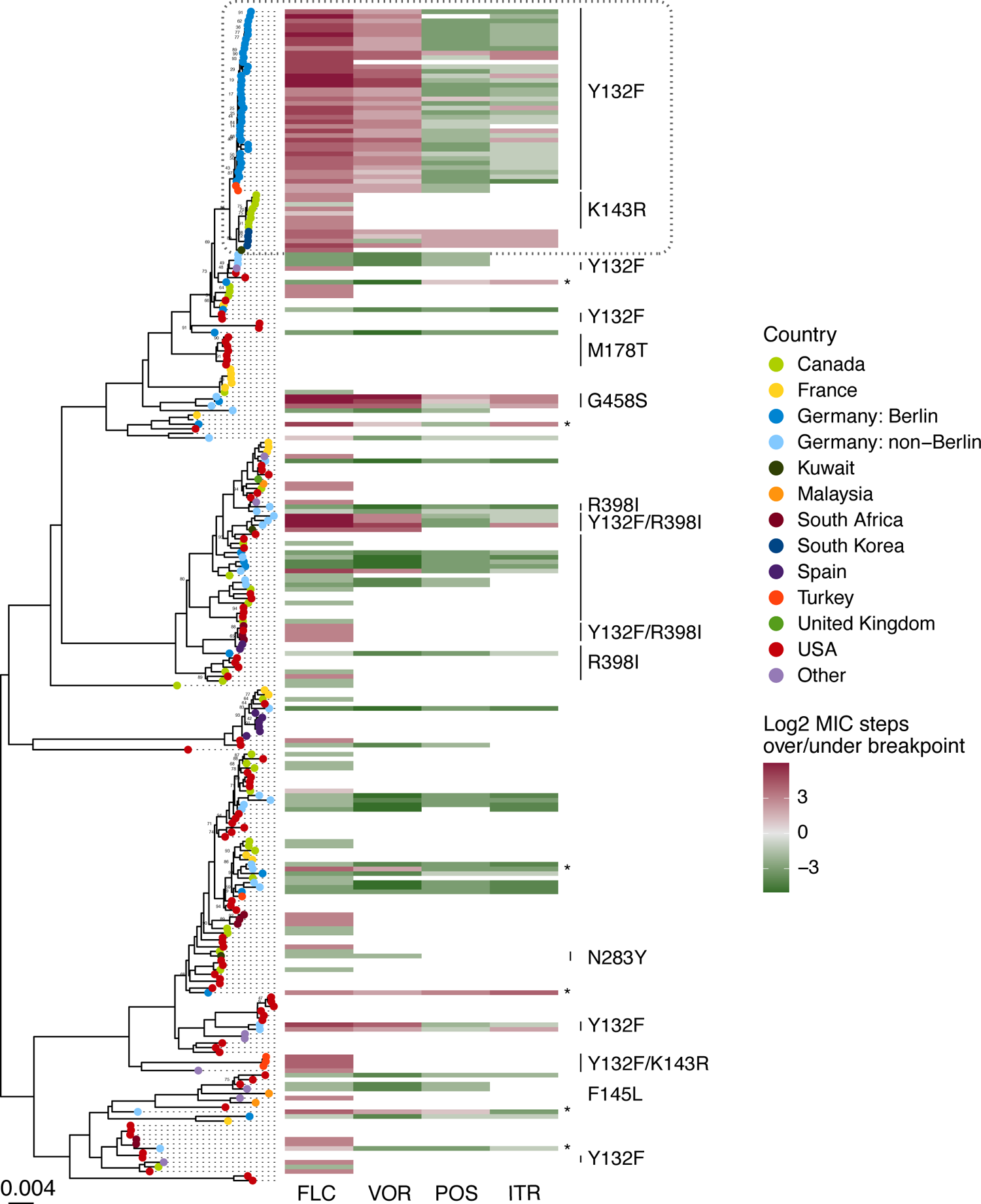
Whole genome phylogeny of *Candida parapsilosis* isolates from a global distribution and their azole susceptibility profiles and *ERG11* polymorphisms. Maximum-likelihood phylogeny constructed from SNV data from 51,494 variable positions. Tree is shown rooted at the midpoint. To increase tree readability, samples from the same sequencing study that showed little genetic differentiation were randomly downsampled to a maximum of 6 samples per sequencing project - with the exception of the samples described for the first time in our study. Branches are supported by ultrafast bootstrap values of >0.95 unless otherwise indicated. Where available, azole susceptibility data relative to EUCAST breakpoints and *ERG11* polymorphisms are indicated. The gray dashed box at the top of the phylogeny indicates the Berlin outbreak samples (medium blue) and the closely related samples from Turkey (red-orange), Canada (light green), South Korea (dark blue), and Kuwait (dark green) found in public data. Samples that were sequenced in our study and were resistant to one or more azole, but where no polymorphisms in *ERG11* were detected, are marked with an *.

The quasi-clonal relationship between outbreak isolates was further confirmed by pairwise single nucleotide variant (SNV) analysis, where isolates in the outbreak clade differed by an average of 36±20 SNVs (SD) (Figure 3). In contrast, the mean number of SNVs between outbreak samples and non-outbreak isolates from other locations in Germany was significantly higher at 2,112± 828 SNVs SD (p < 0.0001, Mann-Whitney U test). Further, the number of SNVs observed between duplicate sequencing of the same freezer stock of an isolate was 15, indicating that the number of SNV differences between repeat sequencing of the same sample was similar to the genetic differentiation between outbreak isolates. We did not detect an obvious spatial-genetic relationship among outbreak isolates but did observe some evidence of clock-like evolution (appendix 1 p 7, Spearman R = 0.36, p = 0.063).

**Figure 3.**
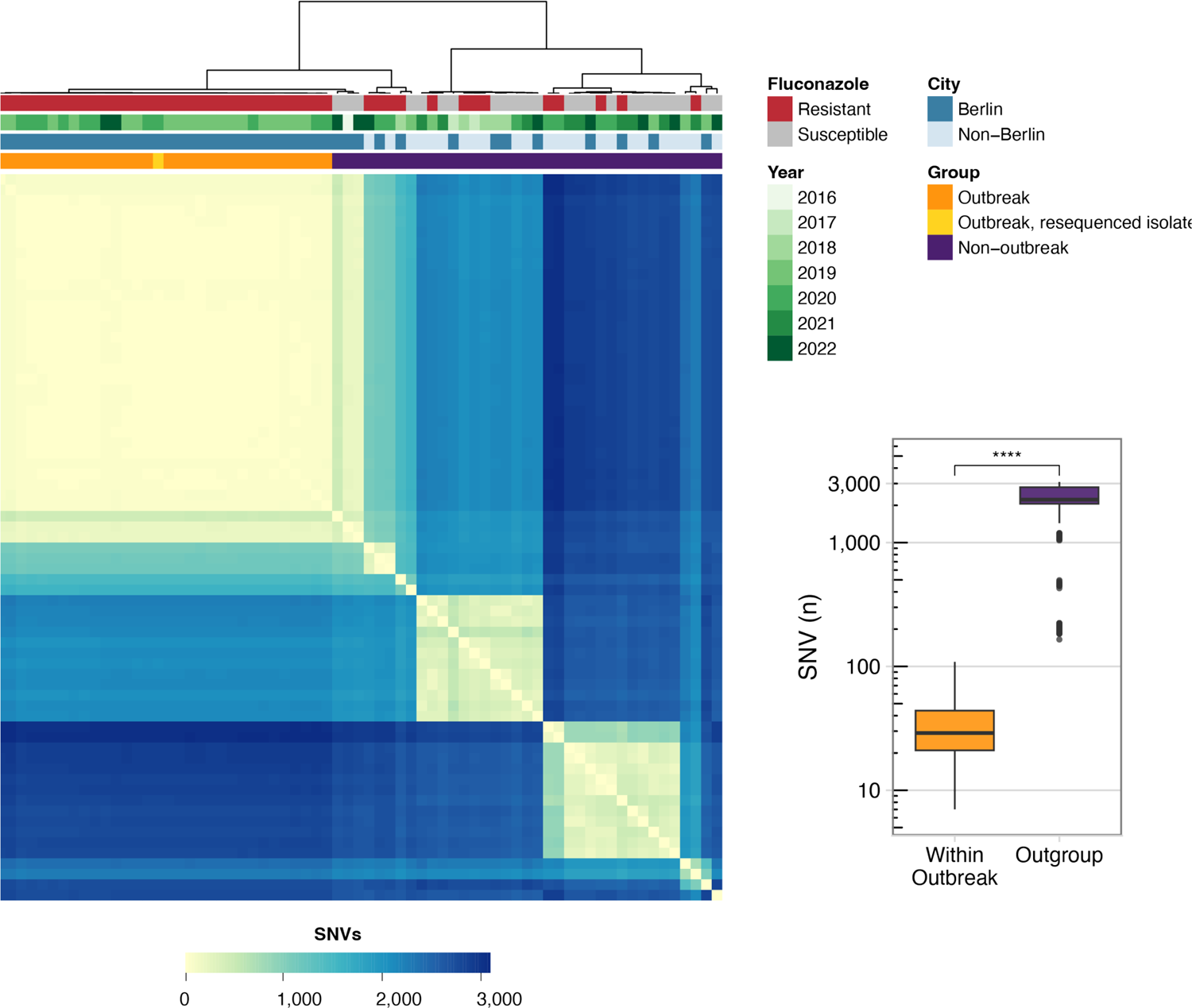
SNV matrix of the German isolates sequenced in this study. Heatmap showing the pairwise SNV differences between the novel German isolates described in this study. Metadata bands indicate (from top to bottom): fluconazole susceptibility, year of sample isolation, and whether the sample is from the city of Berlin (including both outbreak and non-outbreak strains) or another city in Germany. Boxplot shows the number of pairwise SNV differences when the 33 outbreak strains are compared against each other compared to their pairwise differences with the other German isolates sequenced in this study (outgroup). Significance determined by Mann-Whitney U test, **** indicates p < 0.0001.

Strikingly, samples from Turkey^19^, Canada^13^, South Korea^20^, and Kuwait^21^ were closely related to the Berlin outbreak strains (Figure 2). The Canadian isolates were reported as part of a drug resistant outbreak, revealing that this lineage has caused outbreaks on multiple continents. Although closely related, the strains from each geographic region could still be distinguished by pairwise SNV differences and were more similar to each other than to the other regions (appendix 1 p 8). Interestingly, despite their close genetic relationship and all strains being fluconazole resistant, multiple resistance mechanisms were present. The strains from Berlin and Turkey carried an *ERG11* Y132F mutation, while strains from Canada were *ERG11* K143R (another mutation associated with fluconazole resistance) and those from South Korea were *ERG11* wild-type. Altogether, whole genome sequencing confirmed the close genetic relationship of the outbreak strains and the presence of this lineage in multiple other countries.

*ERG11* Y132F was the dominant azole resistant mechanism across all German isolates, with 82% of resistant isolates possessing this mutation (n = 38/38 outbreak; n = 5/13 non-outbreak). Interestingly, the non-outbreak Y132F strains were in two distantly related groups of samples based on the phylogeny (Figure 2; appendix 1 p 9). Each of these Y132F phylogroups contained isolates from geographically distant regions, suggesting that there are at least 3 distinct lineages of Y132F *C. parapsilosis* circulating widely in Germany. We also observed three closely related but unlinked samples from different cities in Germany that were pan-azole resistant and carried a G458S substitution in *ERG11* (Figure 2; appendix 1 p 9) that has been linked to pan-azole resistance previously ^7,22,23^. Also included in our dataset were six newly-sequenced isolates that were resistant to one or more azoles, but did not contain any resistance-associated polymorphisms in *ERG11*. These isolates contained sequence polymorphisms in several genes that have been linked to drug resistance, including the efflux pumps *MDR1* and *CDR1B*, the multidrug resistance gene regulator *MRR1,* and the transcriptional activator *TAC1* (appendix 1 pp 12-13). While a causal role for most of these polymorphisms in drug resistance have not been experimentally established, one resistant isolate harbored a L518F substitution in *TAC1* experimentally proven to confer azole resistance *in vitro* ^24^.

Our analysis to select loci suitable for Sanger sequence-based typing identified four 750 bp loci in CPAR2_101400, CPAR2_101470, CPAR2_108720, and CPAR2_808110 (Table 2). These loci the 386 isolates into 62 unique sequence types (appendix 3) with a Hunter’s Discriminatory Power of 0·93. Discriminatory power assumes unrelated strains, while our dataset contains multiple outbreaks and sets of serial isolates. When the global dataset was pruned as described in appendix 1 p3, the discriminatory power rose to 0·98. Conversely, *C. parapsilosis* is a diploid organism and an appreciable fraction of the discriminatory power comes from whether sequence polymorphisms were heterozygous or homozygous. When site zygosity was not considered, we were only able to identify 43 sequence types across the 386 samples and the discriminatory power was dropped to 0·90. The four selected loci showed a high degree of agreement with the whole genome phylogeny and were able to correctly identify samples to their whole genome-defined genetic cluster with 97·1% sensitivity (0.951-0.982 95% CI) and 99·8% specificity (0.997-0.99 95% CI) (appendix 1 p 6). To confirm that the selected loci were present as a single copy and the discriminatory power was not being inflated by the presence of multiple copies in the genome, we assessed them for genome-wide copy number changes. We did not detect any CNV changes at these loci, either additional copies or loss of the region (appendix 1 p 10).

We validated our *in silico* typing using the primers in Table 2 and a representative 13 samples from our dataset, including four outbreak strains expected to have identical sequence type profiles. In each sample, we detected all anticipated variants by Sanger sequencing and did not discover any variants that were not in the whole genome data (appendix 1 p 14). As expected, the four outbreak samples had identical sequences at the four loci and were assigned to the same sequence type. To facilitate the typing of novel isolates by researchers and clinical labs, FASTA alignments containing representative allelic sequences of the four markers are provided as appendices 4-7.

Finally, we demonstrated the utility of the MLST scheme on seven isolates that were not part of the dataset used to create the typing scheme, but were suspected to be additional outbreak cases based on submitting hospital and antifungal susceptibility profiles (appendix 1 p 4). Their sequence type was identical to the outbreak isolates (appendix 1 p 14), suggesting that they were outbreak cases 26-32 (Figure 1a), which was confirmed by WGS. Taken together, we identified and validated four loci that can be used for sequence-based typing of *C. parapsilosis* and demonstrated their value in rapidly identifying closely related samples.

## Discussion

In this work, we use genomic epidemiology coupled with patient tracking to investigate an outbreak of *ERG11* Y132F azole-resistant *C. parapsilosis* that occurred over several years and medical centers in Berlin, Germany. We use a large-scale genomic dataset composed of 386 *C. parapsilosis* isolates to investigate outbreak dynamics, confirm the high degree of relatedness among outbreak isolates, and compare them to isolates from around the world—revealing that the outbreak clone is closely related to samples from several other continents. Based on the increasing number of outbreaks for *C. parapsilosis* and the need to rapidly discriminate between isolates, we also establish and validate a set of four loci for Sanger sequence-based strain typing.

Due to slow outbreak kinetics and the low case density, the outbreak might have gone unnoticed if not for the atypical susceptibility pattern that was detected by clinicians and clinical microbiologists. The reconstructed transmission dynamics suggest that outbreaks of *C. parapsilosis* may result from the reintroduction of outbreak strains from other hospitals, underlining the challenges of outbreak control and the need for heightened infection control measures. In our case, patient transfers were a common occurrence among cases and facilitated inter-hospital transmission and subsequent intra-hospital transmission. Unfortunately, high-resolution temporal and spatial monitoring of patients is lacking for other outbreaks, so it is unclear the degree to which this has driven other reported *C. parapsilosis* outbreaks. However, the pattern of prolonged but tightly related isolates causing disease across several years and multiple medical centers matches other outbreak reports ^10,23,25,26^, suggesting that reintroductions and/or patient transfers could have also contributed to the prolonged nature of these outbreaks. Though our study period concluded in 2022, given the prolonged and sporadic nature, we believe it is unlikely that the outbreak has stopped for good.

One limitation to our study is that while patients without infection were screened for colonization during the first wave of cases at Hospital #1, comprehensive screening of all the involved hospitals and throughout the entire study period was not performed. This obscures an understanding of the relationship between colonization and subsequent infection during the outbreak and a mechanistic understanding of how *C. parapsilosis* is transmitted within healthcare facilities.

The finding of azole-resistant strains from different continents that were very closely related to the strains spreading within healthcare facilities in Berlin highlights the global challenge of antifungal resistant pathogens and the emergence of *Candida* spp that are easily transmitted patient-to-patient. It is likely that drug resistance was acquired independently and recently in this lineage, given the close genomic relationship but different resistance mechanisms in strains from Germany, Canada, and South Korea. How this lineage acquired drug resistance and how it has spread across continents are open questions, but we hypothesize that it may have enhanced properties that facilitate its transmission and persistence. The observation of highly related clones observed on multiple continents mirrors the behavior of *Candida auris*. Nearly identical clades of this pathogen have been reported on multiple continents and continue to spread globally ^27,28^, underscoring the fact that *C. parapsilosis* has the potential to become a problem of similar scale. Additionally, three phylogenetically distinct Y132F lineages of fluconazole-resistant *C. parapsilosis* were discovered in Germany during the study period, each with multiple isolates reported from distinct geographic areas. We also identified six isolates that are resistant by an unknown mechanism, which will be an interesting area to follow up on using genome-wide association approaches. Taken together these observations demonstrates that fluconazole-resistant *C. parapsilosis* is endemic in Germany, mirroring what has been recently been reported in other countries ^3^, and highlighting the changing epidemiology of the pathogen that clinicians should be aware of.

Microsatellite typing in *C. parapsilosis* is typically performed using four loci and provides a discriminatory power between 0·63 ^29^ and 0·99 ^15^. The four loci MLST established in this study achieved a discriminatory power of 0·93, within the range of what is achieved using microsatellites. Importantly, MLST methodology could unambiguously compare samples from >9 years and around the world, a task not possible using existing microsatellite strategies based on gel electrophoresis profiles. Taken together, our study provides a high-resolution description of an azole-resistant *C. parapsilosis* outbreak, identifies hospital transfers as recurring source of pathogen introductions, and defines a sequence-based typing scheme for epidemiology and rapid identification of potential outbreaks.

### Data sharing

Raw FASTQ files for novel isolates sequenced in this study were uploaded to the NCBI Sequence Read Archive and are publicly available under BioProject PRJNA996760. Accession numbers for the publicly available sequence data used in this study are listed in appendix 2. Genbank records for the Sanger sequencing of the MLST loci performed in this study are listed in appendix 1 p 14. FASTAs containing representative allele sequences for each of the four typing loci are available as appendices 4-7. Most data analysed in this study are included within the study or in the appendices. However, remaining datasets can be made available from the corresponding author on reasonable request.

## Data Availability

Raw FASTQ files for novel isolates sequenced in this study were uploaded to the NCBI Sequence Read Archive and are publicly available under BioProject PRJNA996760. Accession numbers for the publicly available sequence data used in this study are listed in appendix 2. Genbank records for the Sanger sequencing of the MLST loci performed in this study are listed in appendix 1 p 14. FASTAs containing representative allele sequences for each of the four typing loci are available as appendices 4-7.

## Acknowledgements

We are grateful to Peggy Karanatsiou of the Friedrich Schiller University and Philipp Hupel, Carmen Karkowski, and Christiane Weigel of the NRZMyk for technical assistance. We thank Britta Schweickert of the Robert-Koch-Institute for contributing regional hospital data from the antibiotic consumption surveillance system. We also acknowledge MSD and Pfizer for providing the drugs for antifungal susceptibility testing to the NRZMyk free of charge.

## Contributors

AEB and GW conceptualized the study. PB, FRK, BG, RP, OK, GW and AEB contributed to investigation, methodology, and formal analysis. PB, GW, and AEB were responsible for visualization. AEB did the statistical analysis. AEB, PB, and GW wrote the original draft. All authors participated in reviewing and editing the manuscript. All authors had full access to all the data in the study and accept responsibility for the decision to submit for publication. AEB and GW verified the underlying data of the study.

