## Appendix 1 for "Genomic reconstruction of an azole-resistant *Candida parapsilosis* outbreak and the creation of a multilocus sequence typing scheme: a retrospective observational and genomic epidemiology study"

### Supplemental Methods

Description of types of hospitals and patient care setting impacted by the outbreak

The outbreak affected both ICU and non-ICU patients. All hospitals involved in the outbreak are part of a transferral network providing specialized care for cardiology / cardio-thoracic patients, including a major cardiac surgery center, post-operative intensive, intermediate and standard care as well as rehabilitation. Consequently, patients affected by the outbreak mainly had underlying cardiac diseases, for which treatment frequently involved surgical interventions including cardiac assist devices.

Antifungal susceptibility testing

*In vitro* antifungal susceptibility testing was performed by broth microdilution technique following the European Committee on Antimicrobial Susceptibility Testing (EUCAST) standard methodology for the following antifungals: fluconazole (FLC; Pfizer Inc., Peapack, NJ, USA), itraconazole (ITR, Chemicalpoint, Deisenhofen, Germany), posaconazole (POS; MSD, Rahway, NJ, USA), and voriconazole (VOR; Pfizer Inc., Peapack, NJ, USA). Minimum inhibitory concentration (MICs) were assessed with a nephelometer (Labsystems Nepheloskan Ascent Microplate Reader Type 750, Helsinki, Finland) after 24 h of incubation at 35°C. The endpoint of growth was as a 50% inhibition in comparison with the drug-free control. Reference strain *C. parapsilosis* ATCC 22019 was used as a quality control.

DNA extraction

For genomic DNA extraction, single colonies were grown overnight on YPD plates at 35°C. Fungal material was transferred to a tube containing acid-washed glass beads and lysis buffer (50 mM Tris, 50 mM sodium EDTA, 3% [wt/vol] sodium dodecyl sulfate [SDS] [pH 8]). The samples were homogenized using a vortex adapter, followed by incubation in a thermomixer. Next, cellular debris was pelleted by centrifugation, and the supernatant was transferred to a new tube. RNAse treatment was performed for 1h at 37°C. Subsequently, 3M sodium acetate was added to the tube, samples were incubated at -20°C, then centrifuged and the supernatant transferred to a new tube. DNA was then precipitated using EtOH and the resulting pellet washed twice. DNA extracts that did not meet the requirement of purity were purified using the Monarch genomic DNA purification kit (New England BioLabs Inc., Frankfurt a. M., Germany). Isolates were identified as *C. parapsilosis* prior to WGS by Sanger sequencing of the ITS locus. This locus was amplified with the forward primer V9G (5′-TTACGTCCCTGCCCTTTGTA-3′) and the reverse primer LS266 (5′-GCATTCCCAAACAACTCGACTC-3). The resulting sequences were compared against *C. parapsilosis* sequences in the NCBI database using BLAST.

Quality control, alignment, and variant identification

Raw sequence reads were trimmed for quality using TrimGalore v0.6.6 in paired end mode using the default error rate threshold of 0.1. Sequence data quality was assessed before and after trimming using FastQC v0.11.9. Mapping to the CDC317 reference genome (FungiDB, release 50) was performed using bwa-mem2 v2.2.1 [1]. Duplicates were marked using the MarkDuplicates function of Picard v2.24.0. Alignment and genome coverage statistics were assessed using CollectAlignmentMetrics and CollectWgsMetrics from Picard tools (v2.24.0). Only samples with a mean genome coverage of 10x and >90% of reads mapping to the CDC317 reference genome were included in downstream analyses. Variant calling was performed using Freebayes v1.3.2-dirty. Variants were filtered using vcffilter v0.2 and the following filter string: “QUAL > 30 & DP > 5 & SAF > 0 & SAR > 0 & RPR > 1 & RPL > 1”. Variants were annotated using SnpEff v5.1 [2] and a database made from the CDC317 reference genome files and using the alternative yeast nuclear and yeast mitochondrial codon tables. For defining upstream and downstream regions, a cutoff of 500 bp was used.

Phylogenomic and SNV-based analyses

For the phylogeny and SNV distance-based analyses, only variant positions with non-zero coverage in all samples and SNVs were included. To represent heterozygous sites, SNVs were converted to their corresponding IUPAC base. Pairwise SNV distances between isolates were calculated using SNP Dist [3]. A maximum-likelihood (ML) phylogeny was inferred by IQ-TREE 2 v2.2.0.3 [4] using the GTR+ASC model to account for ascertainment bias as constant positions were excluded. The ModelFinder module of IQ-TREE was used to confirm that GTR was also the best fitting model based on Akaike and Corrected Akaike Information Criterion. The *ERG11* amino acid annotations on the phylogeny are reported relative to the *C. parapsilosis* reference genome CDC317, except for cDNA position 395, corresponding to amino acid position 132. CDC317 is azole resistant and contains a Y132F substitution relative to the EUCAST control strain ATCC 22019. Given this, samples reporting no variant at position 395 of the CDS were re-coded to a Y132F amino acid substitution and samples recording a 395T>A change relative to CDC317 were reported as not having an amino acid substitution at position 132.

For regression analysis, a phylogeny containing only the German outbreak isolates was constructed using the methodology described as above. Root-to-tip distance in the phylogeny was calculated using Clockor2 [5].

Phylogenetic clusters were identified using TreeCluster v1.0.3 [6] using a cut threshold of 0.55. The identified clusters were labeled in accordance with the five major clades described in [7]. TreeCluster calculated sufficient intraclade genetic distance to further subdivide clades 2 and 4. Clusters belonging to these clades were assigned the IDs of 2a, 2b and 4a-i.

Identification of discriminatory loci for sequence typing

Variant profiles of gene coding regions were obtained from the vcf files and analyzed by hierarchical clustering. The top 212 CDS were further evaluated in 750 bp sliding windows with a step size of 50 bp. Windows including intronic sequence were excluded for downstream selection. The top eight most-discriminatory individual loci were combined in all combinations and the discriminatory power and number of sequence types assessed to determine the optimal number of loci for the typing scheme. To ensure that all potential sequence typing loci were present in the genomes of all samples as a single copy, loci were screened for the presence of the presence of copy number variations (CNVs) using Control-FREEC v11.6 [8] (minExpectedGC 0.25, maxExpectedGC 0.45), DELLY v1.1.6 [9], and CNVator v0.4.1 [10]. Multiple tools utilizing different approaches for detecting copy number changes, including tools utilizing read depth, GC normalization, and split read mapping, were used as many CNV calling software suffer from false positives. DELLY and CNVator were run with default parameters. As FREEC uses GC normalization which varies by organism, 0.25 was used for the minExpectedGC and 0.45 for maxExpectedGC. Loci that fell within CNVs detected by two or more callers were excluded from further consideration. Several samples from the publicly available data displayed prominent chromosome-end bias[11]. As this overrepresented coverage at the chromosome ends reduces confidence for CNV calling, these samples were excluded from CNV analysis.

Four loci was chosen over the six to seven used in other MLST schemes because it balanced workload and cost with the diminishing returns of additional loci (see p 10 of this appendix).

Discriminatory power assumes genetically unrelated strains, whereas our dataset contains multiple outbreaks and sets of serial isolates. To calculate discriminatory power using a dataset that did not violate this assumption as strongly, samples that were from the same BioProject and formed a terminal, monophyletic group, were downsampled to a maximum of four, resulting in a pruned dataset of 225 samples.

*In silico* strain typing

*In silico* locus sequence was extracted from vcf files using the consensus function of BCFtools v1.9 [12] and aligned using MUSCLE v3.8. As BCFtools is not able to convert heterozygous sites to their corresponding IUPAC base, this was performed after alignment using an in-house R script. Concatenated alignments from the four loci were built into a maximum-likelihood phylogeny using RaxML (v2.0.10) and the GTR substitution model with 100 bootstrap replicates.

### Supplemental Figure 1

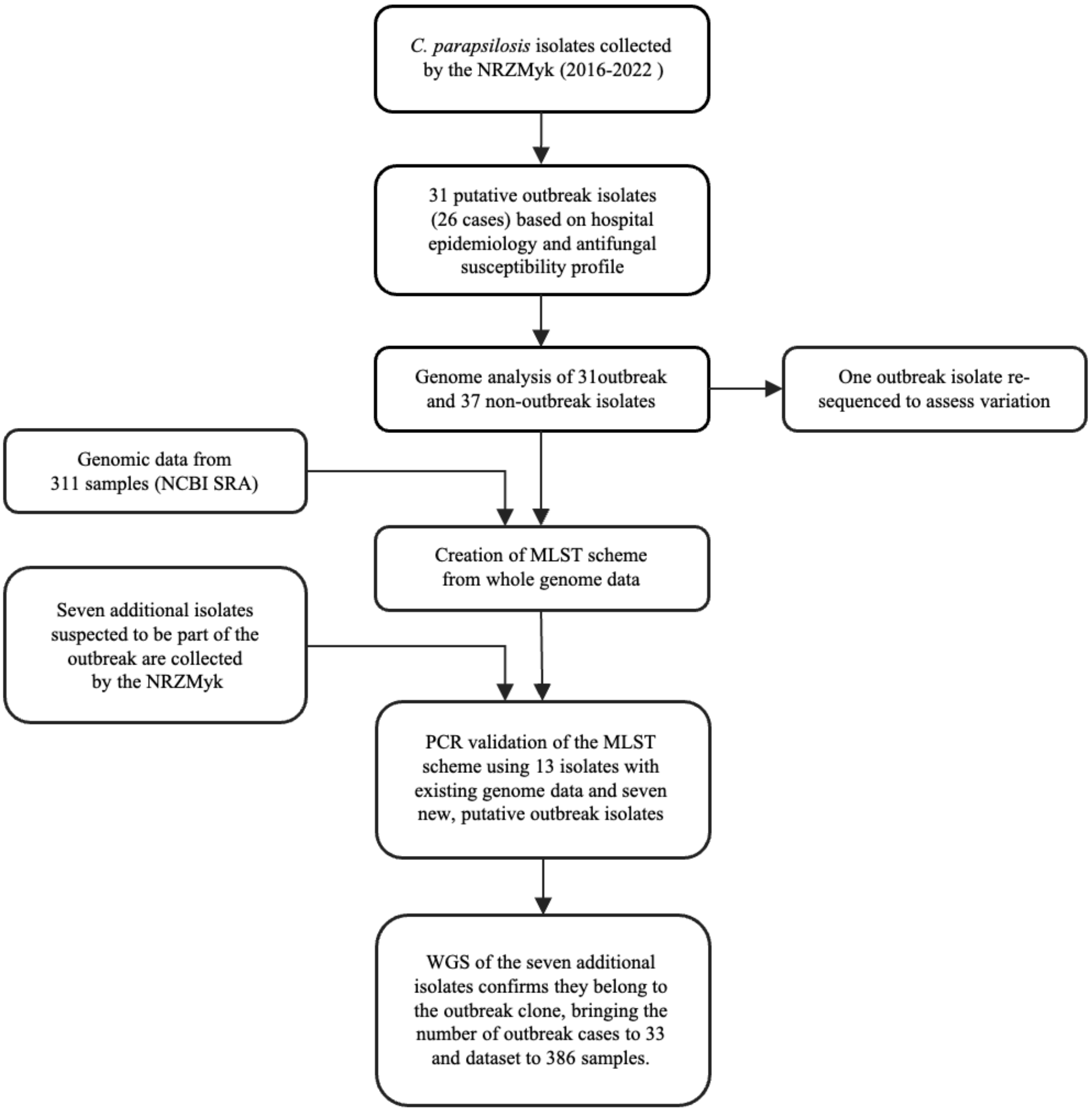

**Flowchart illustrating the samples included and key steps in this study**. The study was performed on a dataset of 386 samples corresponding to 75 newly sequenced isolates and 311 global samples. One outbreak isolate was resequenced to assess biological and technical variation and is not included in the 386 samples and was only included in the analyses of pairwise SNV differences of the German isolates sequenced in this study (Figure 3). The seven additional outbreak isolates, corresponding to outbreak cases 26-33, were initially not part of the whole genome collection and were therefore not included in the construction of the typing scheme, but were included in all genomic analyses *post hoc* once it was determined that their MLST and genome profile matched the outbreak clone.

### Supplemental Figure 2

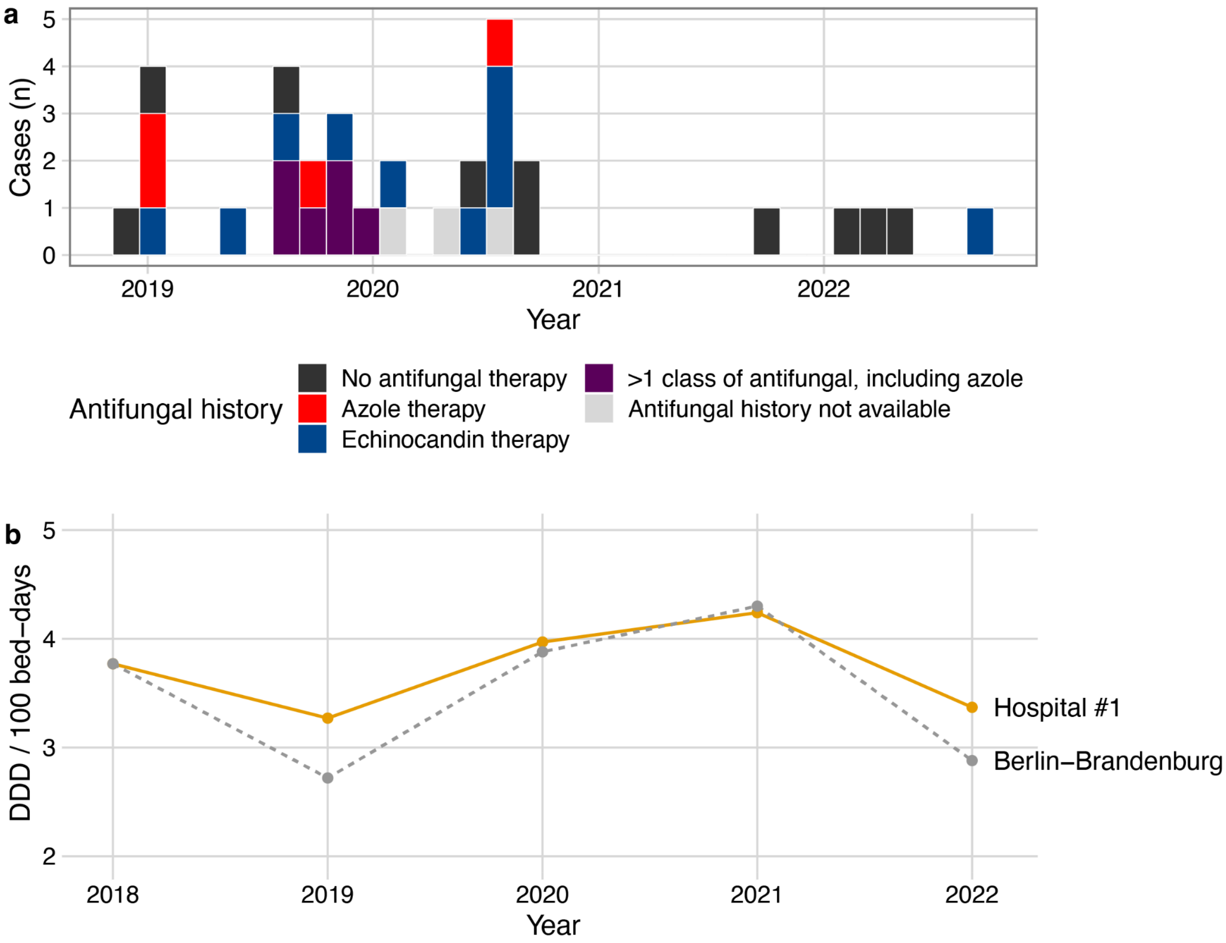

**Prior antifungal treatment in outbreak cases and regional consumption of azoles during the study period.** (a) Prior antifungal therapies for outbreak cases within 60 days prior to isolation of *C. parapsilosis* (b) Defined daily dose (DDD) per 100 bed-days of azoles (J02AC) at Hospital #1 and other hospitals of the same category (care level three) in the combined Berlin and Brandenburg regions (n = 2-4 care centers depending on the year).

### Supplemental Figure 3

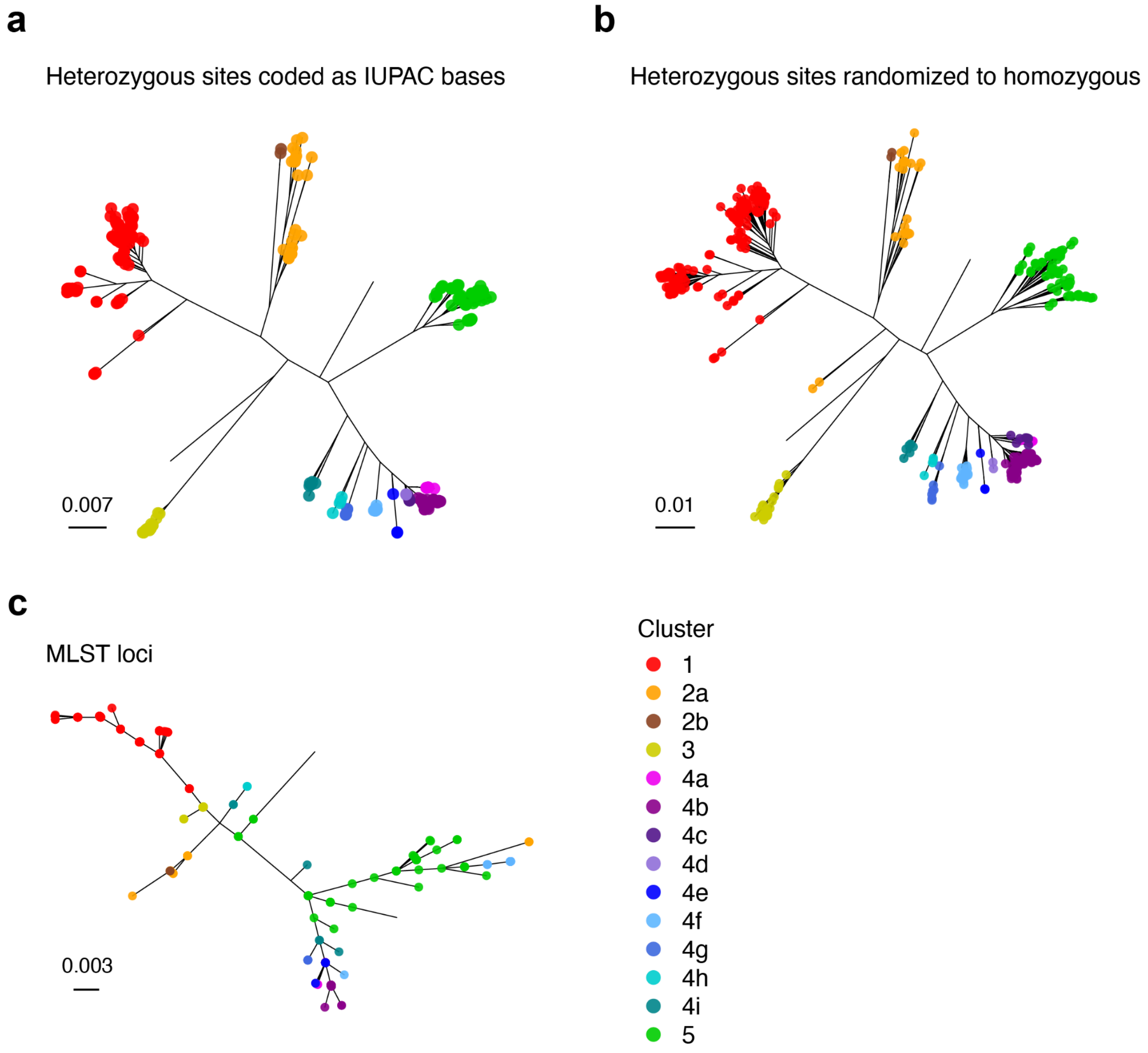

**Impact of heterozygous sites on the whole genome SNV phylogeny.** (a) Maximum-likelihood whole genome SNV phylogeny constructed 51,494 variable positions with heterozygous sites encoded as IUPAC bases. Tip points denote genetic clusters. (b) Maximum-likelihood whole genome SNV phylogeny where heterozygous sites were randomized to homozygous (c) Maximum-likelihood phylogeny constructed using the four MLST loci from CPAR2_101400, CPAR2_101470, CPAR2_108720, and CPAR2_808110, alignment length 2,903 bp. In (b) and (c), tip color indicates the genetic cluster samples were assigned based on the IUPAC-encoded phylogeny in panel (a).

### Supplemental Figure 4

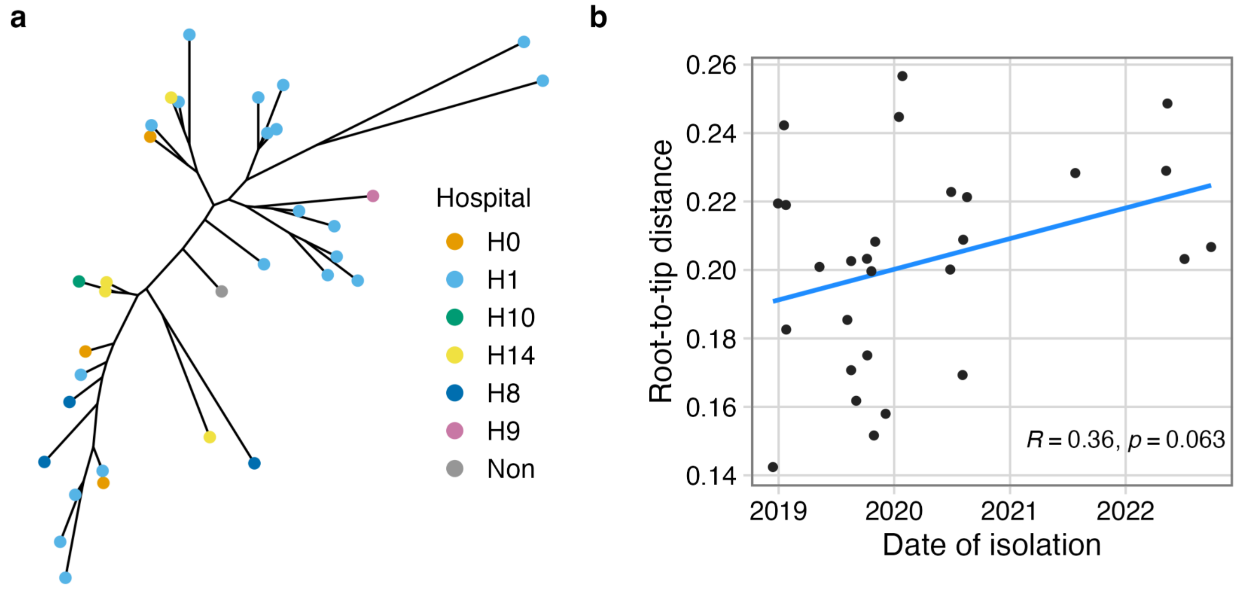

**Genetic relationship vs. temporal and spatial relationships in the outbreak cluster.** (a) Maximum-likelihood phylogeny of the German outbreak cluster. Tip point color indicates the hospital of isolation. A single case was identified in an ambulatory patient, which is indicated with “Non” for the inpatient hospital. (b) Genetic distance is measured as the root-to-tip distance in a phylogeny containing isolates from the German outbreak cluster. Best fitting root for the phylogeny was selected based on residual-mean-squared. Correlation value calculated by Spearman correlation.

### Supplemental Figure 5

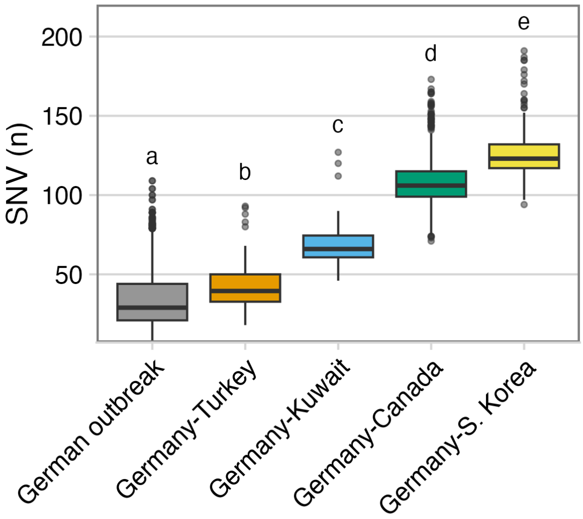

**Pairwise SNV differences between the German outbreak cluster and closely related isolates from other geographic regions**. Pairwise distances calculated among the German outbreak isolates (n = 33 isolates) and these isolates compared to n = 2 from Turkey, n = 1 from Kuwait, n = 13 from Canada, and n = 4 from South Korea. Statistical significance was determined using a one-way analysis of variance and Tukey's honest significance test. The letters denote significance as a compact letter display, where groups that are not significantly different from each other are indicated with the same alphabet letter; $p < 0.05$.

### Supplemental Figure 6

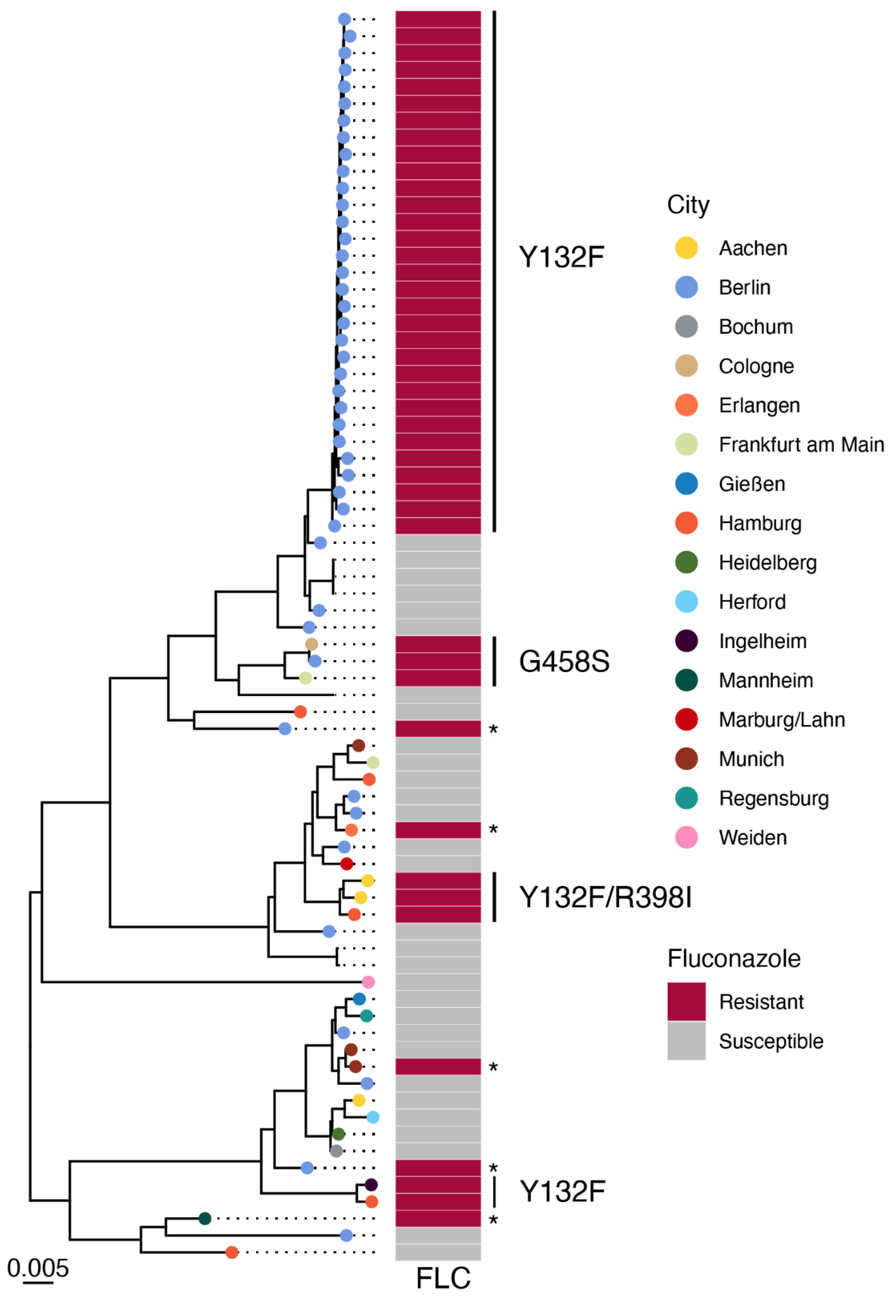

**Whole genome phylogeny of *Candida parapsilosis* isolates from Germany, their fluconazole susceptibility, and *ERG11* polymorphisms**. Maximum-likelihood phylogeny constructed from SNV data from 51,494 variable positions. Tree is shown rooted at the midpoint. Tip colors indicate the city where the isolate originated from. Fluconazole resistant isolates where no polymorphisms in *ERG11* were detected are marked with an asterisk.

### Supplemental Figure 7

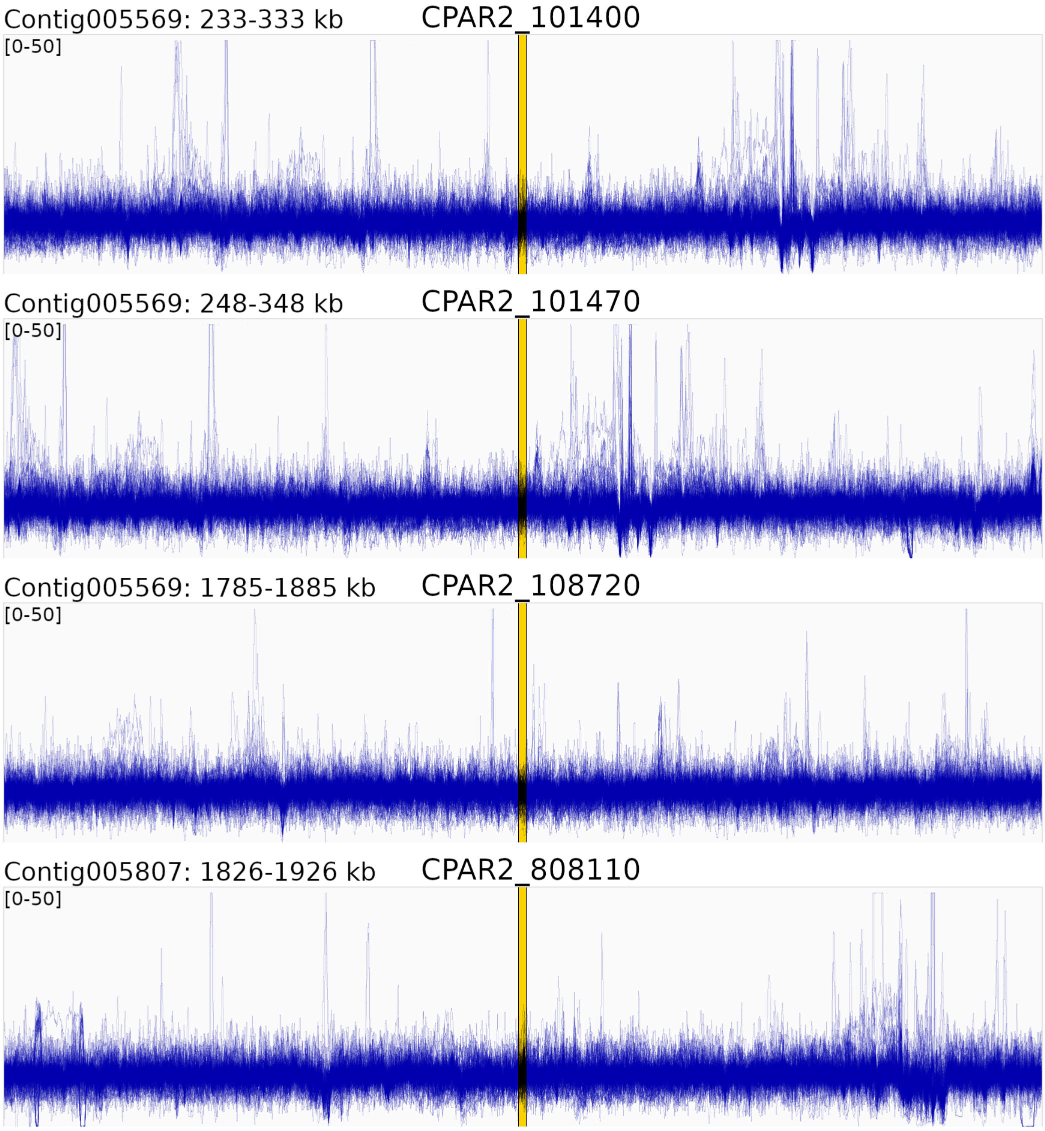

**Genomic coverage at the four sequence typing loci plus 50 kbp of up- and downstream sequence.** Overlaid line traces of normalized read depth. The region used for sequence typing is indicated in the yellow box. Coverage depth was calculated as reads per million (RPM).

### Supplemental Figure 8

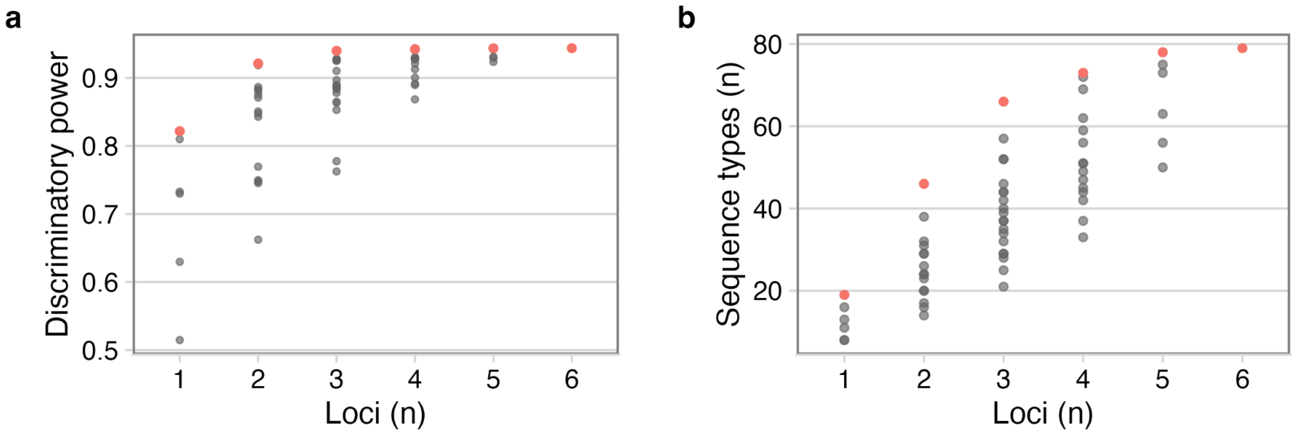

**Relationship between the number of loci, discriminatory power, and number of sequence types.** Using the six most-discriminatory individual loci, all combinations were sampled and the resulting discriminatory power (a) and number of sequence types (b) are shown as points. The best performing combination for each number of loci is highlighted in red.

### Supplemental Table 1.

**Genetic variants observed in resistance-associated genes among azole-resistant isolates.** Variants that were also observed in azole-susceptible isolates are not listed.

| | ***CPAR2_303740 (ERG11)*** | ***CPAR2_304370 (CDR1b)*** | ***CPAR2_301760 (MDR1)*** | ***CPAR2_603010 (MDR1b)*** | ***CPAR2_207540 (MDR1)*** | ***CPAR2_303510 (TAC1)*** | ***CPAR2_807270 (MRR1)*** | ***CPAR2_207280 (UPC2)*** | ***CPAR2_212210 (ALD5)*** | ***CPAR2_401350 (DAG7)*** | ***CPAR2_405010 (ERG6)*** |
|---|---|---|---|---|---|---|---|---|---|---|---|
| **Outbreak** | Y132F | L149Q | | | | | | | | | |
| **2017-535** | Y132F | | | | | | | | | | |
| **2020-561** | Y132F | | | | | | G623D | S392R | | | |
| **2020-315** | G458S | | | | | D444N | | | | | |
| **2022-0454** | G458S | | R303K | | | | | | | | |
| **2016-333** | | P70fs | | | | | | | | | |
| **2018-148** | | A1214V | | F463I | G247R | Y792fs | | | | | |
| **2019-498** | | | | | | | G583E | | | | |
| **2020-725** | | | | | | | | | | | |
| **2021-0166** | | | | | | | S629G | N455D | | P77fs | |
| **2021-0782** | | | | T307K | | | N725Y | | T18N | | C300Y |

Abbreviations in addition to standard one-letter amino acid codes: fs = frameshift; * = nonsense mutation (premature stop codon).

### Supplemental Table 2.

**Minimum inhibitory concentrations (MICs) for the isolates listed in Supplementary Table 1.** MICs are represented as mg/l.

| ***ERG11*** | **Isolate** | **FLC** | **VOR** | **POS** | **ITR** |
|---|---|---:|---:|---:|---:|
| Y132F | Outbreak* | 64 | 1 | 0.03 | 0.125 |
| Y132F | 2017-535 | >64 | 4 | ≤0.016 | 0.5 |
| Y132F | 2020-561 | 64 | 2 | 0.03 | 0.125 |
| G458S | 2020-315 | >64 | 4 | 0.125 | 0.5 |
| G458S | 2022-0454 | 32 | 2 | 0.06 | 0.25 |
| Wildtype | 2016-333 | 0.5 | ≤0.016 | 0.125 | 0.25 |
| | 2018-148 | >64 | 0.25 | 0.03 | 0.5 |
| | 2019-498 | 64 | 1 | ≤0.016 | 0.125 |
| | 2020-725 | 32 | 0.5 | ≤0.016 | 0.03 |
| | 2021-0166 | 16 | 0.5 | 0.5 | 1 |
| | 2021-0782 | 32 | 0.5 | 0.125 | 0.03 |

* Median MIC values; the full MIC range is available in Table 1

### Supplemental Table 3.

***C. parapsilosis*** **isolates used for laboratory validation of sequence typing markers as well as their genetic cluster and sequence type identity and GenBank accession number for the Sanger data generated from each sample.**

| | | GenBank accession | | | |
|---|---|---|---|---|---|
| **Isolate ID** | **Sequence type** | **CPAR2_101400** | **CPAR2_101470** | **CPAR2_108720** | **CPAR2_808110** |
| NRZ-2016-033 | ST 01 (1-1-1-1) | PP719301 | OR413203 | OR413216 | OR413242 |
| NRZ-2018-148 | ST 01 (1-1-1-1) | PP719302 | OR413204 | OR413217 | OR413243 |
| NRZ-2018-167 | ST 03 (3-1-1-2) | PP719303 | OR413205 | OR413218 | OR413244 |
| NRZ-2019-030 | ST 06 (4-1-1-2) | PP719304 | OR413206 | OR413219 | OR413245 |
| NRZ-2019-074 | ST 07 (5-1-1-5) | PP719305 | OR413207 | OR413220 | OR413246 |
| NRZ-2019-077 | ST 01 (1-1-1-1) | PP719306 | OR413208 | OR413221 | OR413247 |
| NRZ-2019-337 | ST 06 (4-1-1-2) | PP719307 | OR413209 | OR413222 | OR413248 |
| NRZ-2019-498 | ST 03 (3-1-1-2) | PP719313 | OR413210 | OR413223 | OR413249 |
| NRZ-2019-532 | ST 01 (1-1-1-1) | PP719308 | OR413211 | OR413224 | OR413250 |
| NRZ-2019-747 | ST 01 (1-1-1-1) | PP719309 | OR413212 | OR413225 | OR413251 |
| NRZ-2020-315 | ST 06 (4-1-1-2) | PP719310 | OR413213 | OR413226 | OR413252 |
| NRZ-2020-540 | ST 01 (1-1-1-1) | PP719311 | OR413214 | OR413227 | OR413253 |
| NRZ-2020-726 | ST 07 (5-1-1-5) | PP719312 | OR413215 | OR413228 | OR413254 |
| NRZ-2021-0591 | ST 08 (1-1-1-6) | PP719314 | OR987460 | OR987469 | OR987451 |
| NRZ-2022-0323 | ST 08 (1-1-1-6) | PP719315 | OR987461 | OR987470 | OR987452 |
| NRZ-2022-0798 | ST 08 (1-1-1-6) | PP719316 | OR987462 | OR987471 | OR987453 |
| NRZ-2023-0851 | ST 08 (1-1-1-6) | PP719318 | OR987454 | OR987463 | OR987445 |
| NRZ-2023-0852 | ST 08 (1-1-1-6) | PP719319 | OR987455 | OR987464 | OR987446 |
| NRZ-2023-0854 | ST 08 (1-1-1-6) | PP719321 | OR987456 | OR987465 | OR987447 |
| NRZ-2023-0850 | ST 08 (1-1-1-6) | PP719317 | OR987457 | OR987466 | OR987448 |
